# Association between Functional Inhibitors of Acid Sphingomyelinase and Reduced Risk of Intubation or Death in Individuals Hospitalized for Severe COVID-19: results from an observational multicenter study

**DOI:** 10.1101/2021.02.22.21252209

**Authors:** Nicolas Hoertel, Marina Sánchez-Rico, Erich Gulbins, Johannes Kornhuber, Alexander Carpinteiro, Eric J. Lenze, Angela M. Reiersen, Miriam Abellán, Pedro de la Muela, Raphaël Vernet, Carlos Blanco, Nathanaël Beeker, Antoine Neuraz, Philip Gorwood, Jesús M. Alvarado, Pierre Meneton, Frédéric Limosin, On behalf of AP-HP / Université de Paris / INSERM COVID-19 research collaboration and AP-HP COVID CDR Initiative

## Abstract

Several medications commonly used for a number of medical conditions share a property of functional inhibition of acid sphingomyelinase (ASM), or FIASMA. Preclinical and clinical evidence suggest that the (ASM)/ceramide system may be central to SARS-CoV-2 infection. We examined the potential usefulness of FIASMA use among patients hospitalized for severe COVID-19 in an observational multicenter retrospective study conducted at Greater Paris University hospitals. Of 2,846 adult patients hospitalized for severe COVID-19, 277 (9.7%) were taking a FIASMA medication at the time of their hospital admission. The primary endpoint was a composite of intubation and/or death. We compared this endpoint between patients taking vs. not taking a FIASMA medication in time-to-event analyses adjusted for sociodemographic characteristics and medical comorbidities. The primary analysis was a Cox regression model with inverse probability weighting (IPW). Over a mean follow-up of 9.2 days (SD=12.5), the primary endpoint occurred in 104 patients (37.5%) who were taking a FIASMA medication, and 1,060 patients (41.4%) who were not. Taking a FIASMA medication was associated with reduced likelihood of intubation or death in both crude (HR=0.71; 95%CI=0.58-0.87; p<0.001) and the primary IPW (HR=0.58; 95%CI=0.46-0.72; p<0.001) analyses. This association remained significant in multiple sensitivity analyses and was not specific to one FIASMA class or medication. These results show the potential importance of the ASM/ceramide system as a treatment target in COVID-19. Double-blind controlled randomized clinical trials of these medications for COVID-19 are needed.

---

Global spread of the novel coronavirus SARS-CoV-2 and its variants, the causative agents of coronavirus disease 2019 (COVID-19), has created an unprecedented infectious disease crisis worldwide.^1–4^ Although the availability of vaccines has raised hope for a decline of the pandemic, the search for an effective treatment for patients with COVID-19 among all available medications is still urgently needed.

Several medications commonly used for a number of medical conditions, such as depression or high blood pressure, are functional inhibitors of acid sphingomyelinase (ASM),^5–9^ or FIASMA (detailed in **Supplementary text 1**). FIASMA medications include certain antidepressants (e.g., fluoxetine, fluvoxamine), antipsychotics (e.g., aripiprazole, chlorpromazine), antihistaminic medications (e.g., clemastine), calcium channel blockers (e.g., amlodipine), and antiarrhythmics (e.g., amiodarone).

Preclinical evidence indicates that SARS-CoV-2 activates the acid sphingomyelinase (ASM)/ceramide system, resulting in the formation of ceramide-enriched membrane domains that facilitate viral entry and infection by clustering ACE2, the cellular receptor of SARS-CoV-2.^5^ An *in vitro* study^5^ showed that several FIASMA^7^ medications, including amitriptyline and fluoxetine, inhibited ASM and the formation of ceramide-enriched membrane domains, and prevented Vero cells from being infected with SARS-CoV-2. Reconstitution of ceramide in cells treated with antidepressant medications having FIASMA properties restored infection with SARS-CoV-2. In healthy volunteers, oral administration of amitriptyline blocked infection of freshly isolated nasal epithelial cells with SARS-CoV-2.^5^ These preclinical data were confirmed by another study that demonstrated an inhibition of the infection of cultured epithelial cells with SARS-CoV-2 by fluoxetine.^10^

Findings from clinical and observational studies are consistent with these preclinical data. First, a randomized double-blind controlled study^11^ showed significant protective effects of the FIASMA antidepressant fluvoxamine (N=80) versus placebo (N=72) on COVID-19 disease progression in outpatients (absolute difference, 8.7% from the survival analysis; log rank p=0.009). Second, an observational multicenter retrospective study using data from Greater Paris University Hospitals showed that use of antidepressants, mostly FIASMA antidepressants, and of the FIASMA hydroxyzine, were significantly associated with reduced mortality in patients hospitalized for COVID-19.^12,13^ Third, retrospective clinical investigations among hospitalized COVID-19 patients either elderly (N=77)^14^ or with hypertension as the only comorbidity^15^ (N=96) showed that the use of amlodipine (a calcium channel blocker^16^ and a FIASMA) may be associated with decreased mortality. Taken together, these results suggest that the ASM/ceramide system might provide a useful framework for better understanding SARS-CoV-2 infection and favouring the possible repurposing of FIASMA medications against COVID-19.

To our knowledge, no clinical study to date has examined the potential usefulness of FIASMA medications as a class in patients hospitalized for COVID-19. Observational studies of patients with COVID-19 taking medications for other disorders can help to decide which treatment should be prioritized for randomized clinical trials and to minimize the risk for patients of being exposed to potentially ineffective or harmful treatments.^12,13,17–19^

We used data from Greater Paris University Hospitals to examine the association between FIASMA medication use and the composite outcome of intubation or death among patients hospitalized for severe COVID-19. Our primary hypothesis was that FIASMA medication use would be associated with reduced risk of intubation or death among patients hospitalized for severe COVID-19 in time-to-event analyses adjusting for sociodemographic characteristics and medical comorbidities. Additional exploratory analyses examined whether this association was specific to certain FIASMA classes (e.g., FIASMA antidepressants) or individual medications (e.g., amlodipine).

## Methods

### Setting and Cohort Assembly

A multicenter cohort study was conducted at 36 AP-HP hospitals from the beginning of the epidemic in France, i.e. January 24^th^, until May 1^st^, 2020. We included all adults aged 18 years or over who have been hospitalized in these medical centers for severe COVID-19. COVID-19 was ascertained by a positive reverse-transcriptase–polymerase-chain-reaction (RT-PCR) test on nasopharyngeal or oropharyngeal swab specimens. Severe COVID-19 was defined as having at least one of the following criteria at hospital admission : respiratory rate > 24 breaths/min or < 12 breaths/min, resting peripheral capillary oxygen saturation in ambient air < 90%, temperature > 40°C, systolic blood pressure < 100 mm Hg, or high lactate levels>2mmol/L.^20–22^

This observational study using routinely collected data received approval from the Institutional Review Board of the AP-HP clinical data warehouse (decision CSE-20-20_COVID19, IRB00011591, April 8^th^, 2020). AP-HP clinical Data Warehouse initiatives ensure patient information and informed consent regarding the different approved studies through a transparency portal in accordance with European Regulation on data protection and authorization n°1980120 from National Commission for Information Technology and Civil Liberties (CNIL). All procedures related to this work adhered to the ethical standards of the relevant national and institutional committees on human experimentation and with the Helsinki Declaration of 1975, as revised in 2008.

### Data sources

AP-HP Health Data Warehouse (‘Entrepôt de Données de Santé (EDS)’) contains all available clinical data on all inpatient visits for COVID-19 to 36 Greater Paris university hospitals. The data included patient demographic characteristics, vital signs, laboratory test and RT-PCR test results, medication administration data, medication lists during current and past hospitalizations in AP-HP hospitals, current diagnoses, discharge disposition, and death certificates.

### Variables assessed

We obtained the following data for each patient at the time of the hospitalization through electronic health records:^23,24^ sex, age, hospital, obesity, self-reported current smoking status, any medication prescribed according to compassionate use or as part of a clinical trial (e.g. hydroxychloroquine, azithromycin, remdesivir, tocilizumab, sarilumab, or dexamethasone), and medical comorbidities based on ICD-10 diagnosis codes during the visit. These variables are detailed in **Supplementary text 2**.

### Medications with functional inhibition effect on acid sphingomyelinase (ASM)

FIASMA medications were defined as having a substantial *in vitro* functional inhibition effect on ASM (i.e., a residual ASM activity lower than 50%), as detailed elsewhere,^5–9^ and were divided into the following classes according to their Anatomical Therapeutic Chemical (ATC) code:^25^ FIASMA alimentary tract and metabolism medications (e.g., loperamide); cardiovascular system medications, subdivided into calcium channel blockers (e.g., amlodipine) and other cardiovascular medications (e.g., carvedilol); nervous system medications, subdivided according to ATC codes into psychoanaleptic (e.g., amitriptyline) and psycholeptic medications (e.g., chlorpromazine); and respiratory system medications (e.g., desloratadine).

FIASMA medication use was defined as receiving at least one FIASMA medication within the first 24 hours of hospital admission. To minimize potential confounding effects of late prescription of FIASMA medications, patients who initiated a FIASMA medication more than 24 hours after hospital admission were excluded from the analyses. Patients who received at study baseline an antipsychotic or a benzodiazepine while being hospitalized in an intensive care unit (ICU), possibly as an aid to oral intubation, were also excluded.

### Primary endpoint

Study baseline was defined as the date of hospital admission for COVID-19. The primary endpoint was the occurrence of intubation and/or death. For patients who died after intubation, the timing of the primary endpoint was defined as the time of intubation. Patients without an end-point event had their data censored on May 1^st^, 2020.

### Statistical analysis

We calculated frequencies of all baseline characteristics described above in patients receiving or not receiving a FIASMA medication and compared them using standardized mean differences (SMD).

To examine the association between FIASMA medication use at baseline and the endpoint of intubation or death, we performed Cox proportional-hazards regression models ^26^. To help account for the nonrandomized prescription of medications and reduce the effects of confounders, the primary analysis used propensity score analysis with inverse probability weighting (IPW).^27,28^ The individual propensities for receiving a FIASMA medication at baseline were estimated using a multivariable logistic regression model that included sex, age, hospital, obesity, current smoking status, and medical conditions. In the inverse-probability-weighted analyses, the predicted probabilities from the propensity-score models were used to calculate the stabilized inverse-probability-weighting weights.^27^ The association between FIASMA medication use and the endpoint was then estimated using an IPW Cox regression model. In case of non-balanced covariates, an IPW multivariable Cox regression model adjusting for the non-balanced covariates was also performed. Kaplan-Meier curves were performed using the inverse-probability-weighting weights^29,30^ and their pointwise 95% confidence intervals were estimated using the nonparametric bootstrap method.^30,31^

We conducted two sensitivity analyses. First, we performed a multivariable Cox regression model including as covariates the same variables used in the IPW analysis. Second, we used a univariate Cox regression model in a matched analytic sample using a 1:1 ratio, based on the same variables used for the IPW analysis and the multivariable Cox regression analysis. To reduce the effects of confounding, optimal matching was used in order to obtain the smallest average absolute distance across all clinical characteristics between exposed patients and non-exposed matched controls.^32^

We performed four additional exploratory analyses. First, we examined the relationships between each FIASMA class and each individual FIASMA medication with the composite endpoint. Second, we reproduced these analyses while comparing patients receiving any FIASMA medication at baseline to those receiving paracetamol at baseline, to mimic an active comparator. Third, because of discrepancies in the potential FIASMA *in vitro* effect of venlafaxine, mirtazapine, and citalopram,^5,7^ we reproduced the main analyses while considering these molecules as FIASMAs. Finally, we reproduced the main analyses among all patients hospitalized for COVID-19, with and without clinical severity criteria at baseline.

For all associations, we performed residual analyses to assess the fit of the data, check assumptions, including proportional hazards assumption using proportional hazards tests and diagnostics based on weighted residuals,^26,33^ and examined the potential influence of outliers. To improve the quality of result reporting, we followed the recommendations of The Strengthening the Reporting of Observational Studies in Epidemiology (STROBE) Initiative.^31^ Because our main hypothesis focused on the association between FIASMA medication use at baseline and the composite endpoint of intubation or death, statistical significance was fixed *a priori* at two-sided p-value <0.05. Only if a significant association was found, we planned to perform additional exploratory analyses as described above. All analyses were conducted in R software version 3.6.3 (R Project for Statistical Computing).

## Results

### Characteristics of the cohort

Of the 17,131 patients hospitalized for COVID-19, ascertained with a positive COVID-19 RT-PCR test, 1,963 patients (11.5%) were excluded because of missing data or young age (i.e. less than 18 years old of age). Of these 15,168 patients, 3,224 (21.3%) met criteria for severe COVID-19 at hospital admission. Of these 3,224 patients, 378 (11.7%) were excluded because they started a FIASMA medication more than 24 hours after hospital admission (N=343) or because they initiated an antipsychotic or a benzodiazepine in an ICU, possibly as an aid for intubation (N=35). Of the remaining 2,846 adult patients, 277 (9.7%) received a FIASMA medication within the first 24 hours of hospitalization (**Figure 1**), with a mean delay between hospital admission and first FIASMA medication prescription of 0.17 days (SD=0.38).

**Figure 1.**
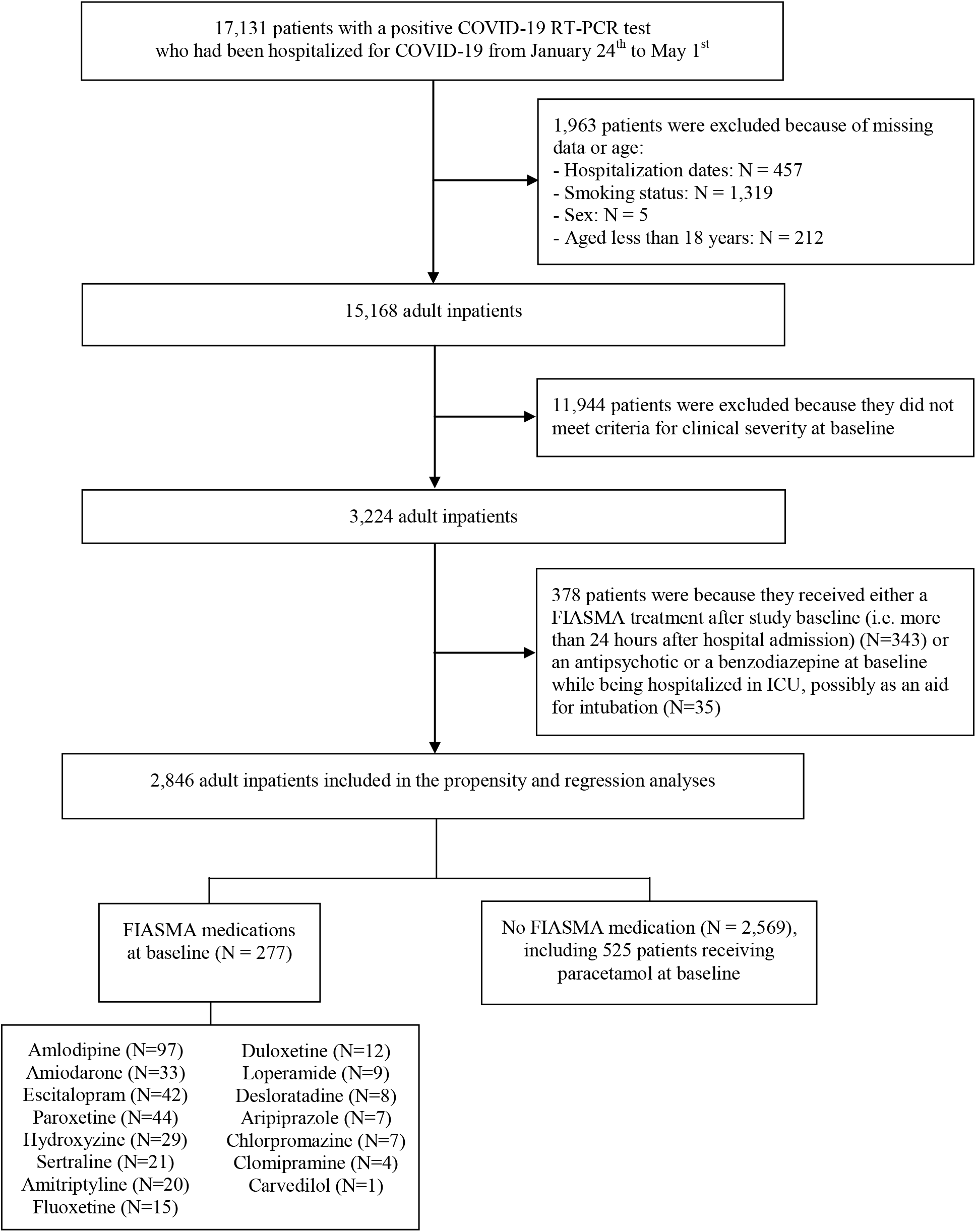
Study cohort.

**Figure 2.**
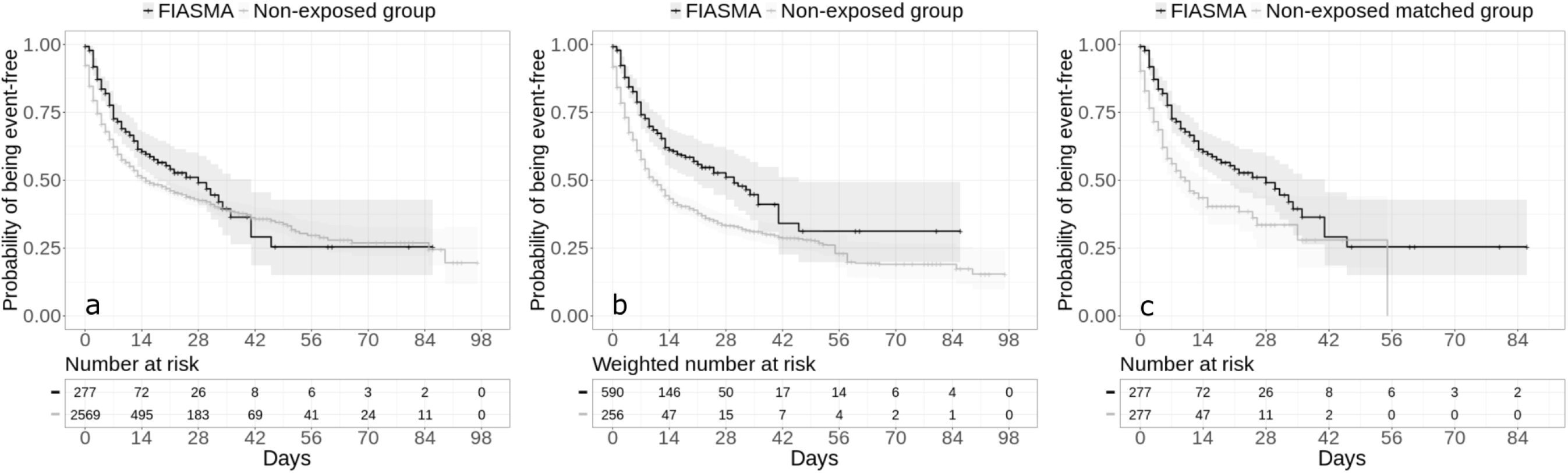
Kaplan-Meier curves for the composite endpoint of intubation or death in the full sample crude analysis (N=2846) (a), in the full sample analysis with IPW (N=2846) (b), and in the matched analytic sample using a 1:1 ratio (N=554) (c) among patients hospitalized for severe COVID-19, according to FIASMA medication use at baseline. The shaded areas represent pointwise 95% confidence intervals. Abbreviations: IPW, inverse probability weighting; FIASMA, Functional Inhibitors of Acid Sphingomyelinase Activity.

RT-PCR test results were obtained after a median delay of 0.9 days (SD=9.4) from hospital admission date. This median delay was similar (i.e., 0.9 days) in the exposed (SD=11.4) and non-exposed (SD=9.3) groups. Over a mean follow-up of 9.2 days (SD=12.5; median=6 days), 1,168 patients (41.0%) had an end-point event at the time of data cutoff on May 1^st^, 2020. Among patients who received a FIASMA medication at baseline, the mean follow-up was 12.0 days (SD=12.9, median=8 days), while it was of 8.9 days (SD=12.4, median=5 days) in those who did not.

All patient characteristics, except current smoking status, diseases of the musculoskeletal system, and Eye-Ear-Nose-Throat disorders were significantly associated with the endpoint. A multivariable Cox regression model showed that sex, hospital in which the patient was treated, obesity, medications according to compassionate use or as part of a clinical trial, cardiovascular disorders, respiratory disorders, neoplasms and diseases of the blood, other infectious diseases, and diseases of the genitourinary system were significantly and independently associated with the endpoint (**Table 1**).

**Table 1.** Characteristics of patients with severe COVID-19 receiving or not receiving a FIASMA medication at baseline (N=2846).

|  | Exposed to any<br>FIASMA<br>medication<br>(N=277) | Not exposed to<br>FIASMA<br>medication<br>(N=2569) | Non-exposed<br>matched group<br>(N=277) | Exposed to any<br>FIASMA<br>medication<br>vs.<br>Not exposed | Exposed to any<br>FIASMA<br>medication<br>vs.<br>Not exposed | Exposed to any<br>FIASMA<br>medication<br>vs.<br>Non-exposed<br>matched group |
| --- | --- | --- | --- | --- | --- | --- |
|  |  |  |  | Crude analysis | Analysis weighted<br>by inverse-probability-weighting weights | Matched analytic<br>sample analysis<br>using a 1:1 ratio |
|  | N (%) | N (%) | N (%) | SMD | SMD | SMD |
| Age |  |  |  | **0.369** | 0.097 | 0.095 |
| 18 to 50 years | 29 (10.5%) | 493 (19.2%) | 29 (10.5%) |  |  |  |
| 51 to 70 years | 88 (31.8%) | 1027 (40.0%) | 99 (35.7%) |  |  |  |
| 71 to 80 years | 63 (22.7%) | 457 (17.8%) | 55 (19.9%) |  |  |  |
| More than 80 years | 97 (35.0%) | 592 (23.0%) | 94 (33.9%) |  |  |  |
| Sex |  |  |  | **0.224** | 0.034 | 0.094 |
| Women | 131 (47.3%) | 933 (36.3%) | 118 (42.6%) |  |  |  |
| Men | 146 (52.7%) | 1636 (63.7%) | 159 (57.4%) |  |  |  |
| Hospital |  |  |  | **0.171** | 0.087 | 0.097 |
| AP-HP Centre - Paris University,<br>Henri Mondor University Hospitals<br>and at home hospitalization | 62 (22.4%) | 660 (25.7%) | 70 (25.3%) |  |  |  |
| AP-HP Nord and Hôpitaux<br>Universitaires Paris Seine-Saint-Denis | 76 (27.4%) | 813 (31.6%) | 68 (24.5%) |  |  |  |
| AP-HP Paris Saclay University | 63 (22.7%) | 561 (21.8%) | 68 (24.5%) |  |  |  |
| AP-HP Sorbonne University | 76 (27.4%) | 535 (20.8%) | 71 (25.6%) |  |  |  |
| Obesity <sup>a</sup> |  |  |  | **0.100** | 0.008 | 0.034 |
| Yes | 67 (24.2%) | 515 (20.0%) | 63 (22.7%) |  |  |  |
| *No* | 210 (75.8%) | 2054 (80.0%) | 214 (77.3%) |  |  |  |
| Smoking <sup>b</sup> |  |  |  | **0.130** | 0.025 | 0.060 |
| *Yes* | 46 (16.6%) | 310 (12.1%) | 40 (14.4%) |  |  |  |
| *No* | 231 (83.4%) | 2259 (87.9%) | 237 (85.6%) |  |  |  |
| Medication according to compassionate use or as part of a clinical trial <sup>c</sup> |  |  |  | 0.040 | 0.020 | 0.024 |
| *Yes* | 83 (30.0%) | 723 (28.1%) | 80 (28.9%) |  |  |  |
| *No* | 194 (70.0%) | 1846 (71.9%) | 197 (71.1%) |  |  |  |
| Other infectious diseases <sup>d</sup> |  |  |  | **0.225** | 0.040 | 0.046 |
| *Yes* | 55 (19.9%) | 301 (11.7%) | 50 (18.1%) |  |  |  |
| *No* | 222 (80.1%) | 2268 (88.3%) | 227 (81.9%) |  |  |  |
| Neoplasms and diseases of the blood <sup>e</sup> |  |  |  | **0.150** | 0.055 | 0.050 |
| *Yes* | 46 (16.6%) | 293 (11.4%) | 41 (14.8%) |  |  |  |
| *No* | 231 (83.4%) | 2276 (88.6%) | 236 (85.2%) |  |  |  |
| Mental disorders <sup>f</sup> |  |  |  | **0.392** | 0.061 | 0.085 |
| *Yes* | 70 (25.3%) | 270 (10.5%) | 60 (21.7%) |  |  |  |
| *No* | 207 (74.7%) | 2299 (89.5%) | 217 (78.3%) |  |  |  |
| Diseases of the nervous system <sup>g</sup> |  |  |  | **0.242** | 0.047 | 0.078 |
| *Yes* | 49 (17.7%) | 243 (9.5%) | 41 (14.8%) |  |  |  |
| *No* | 228 (82.3%) | 2326 (90.5%) | 236 (85.2%) |  |  |  |
| Cardiovascular disorders <sup>h</sup> |  |  |  | **0.392** | 0.099 | 0.022 |
| *Yes* | 147 (53.1%) | 873 (34.0%) | 144 (52.0%) |  |  |  |
| *No* | 130 (46.9%) | 1696 (66.0%) | 133 (48.0%) |  |  |  |
| Respiratory disorders <sup>i</sup> |  |  |  | **0.246** | 0.030 | 0.064 |
| *Yes* | 195 (70.4%) | 1509 (58.7%) | 203 (73.3%) |  |  |  |
| *No* | 82 (29.6%) | 1060 (41.3%) | 74 (26.7%) |  |  |  |
| Digestive disorders <sup>j</sup> |  |  |  | **0.244** | 0.015 | 0.073 |
| *Yes* | 42 (15.2%) | 192 (7.5%) | 35 (12.6%) |  |  |  |
| *No* | 235 (84.8%) | 2377 (92.5%) | 242 (87.4%) |  |  |  |
| Dermatological disorders <sup>k</sup> |  |  |  | **0.152** | 0.010 | <0.001 |
| *Yes* | 14 (5.1%) | 57 (2.2%) | 14 (5.1%) |  |  |  |
| *No* | 263 (94.9%) | 2512 (97.8%) | 263 (94.9%) |  |  |  |
| Diseases of the musculoskeletal system <sup>l</sup> |  |  |  | **0.133** | 0.027 | <0.001 |
| *Yes* | 22 (7.9%) | 121 (4.7%) | 22 (7.94%) |  |  |  |
| *No* | 255 (92.1%) | 2448 (95.3%) | 255 (92.1%) |  |  |  |
| Diseases of the genitourinary system <sup>m</sup> |  |  |  | **0.344** | 0.055 | 0.049 |
| *Yes* | 76 (27.4%) | 353 (13.7%) | 70 (25.3%) |  |  |  |
| *No* | 201 (72.6%) | 2216 (86.3%) | 207 (74.7%) |  |  |  |
| Endocrine disorders <sup>n</sup> |  |  |  | **0.266** | 0.037 | 0.007 |
| *Yes* | 134 (48.4%) | 908 (35.3%) | 135 (48.7%) |  |  |  |
| *No* | 143 (51.6%) | 1661 (64.7%) | 142 (51.3%) |  |  |  |
| Eye-Ear-Nose-Throat disorders <sup>o</sup> |  |  |  | **0.145** | 0.008 | <0.001 |
| *Yes* | 12 (4.3%) | 47 (1.8%) | 12 (4.33%) |  |  |  |
| *No* | 265 (95.7%) | 2522 (98.2%) | 265 (95.7%) |  |  |  |

<sup>a</sup> Defined as having a body-mass index higher than 30 kg/m<sup>2</sup> or an International Statistical Classification of Diseases and Related Health Problems (ICD-10) diagnosis code for obesity (E66.0, E66.1, E66.2, E66.8, E66.9).
<sup>b</sup> Current Smoking status was self-reported.
<sup>c</sup> Any medication prescribed as part of a clinical trial or according to compassionate use (e.g., hydroxychloroquine, azithromycin, remdesivir, tocilizumab, sarilumab or dexamethasone).
<sup>d</sup> Assessed using ICD-10 diagnosis codes for certain infectious and parasitic diseases (A00-B99).
<sup>e</sup> Assessed using ICD-10 diagnosis codes for neoplasms (C00-D49) and diseases of the blood and blood-forming organs and certain disorders involving the immune mechanism (D50-D89).
<sup>f</sup> Assessed using ICD-10 diagnosis codes for mental, behavioural and neurodevelopmental disorders (F01-F99).
<sup>g</sup> Assessed using ICD-10 diagnosis codes for diseases of the nervous system (G00-G99).
<sup>h</sup> Assessed using ICD-10 diagnosis codes for diseases of the circulatory system (I00-I99).
<sup>i</sup> Assessed using ICD-10 diagnosis codes for diseases of the respiratory system (J00-J99).
<sup>j</sup> Assessed using ICD-10 diagnosis codes for diseases of the digestive system (K00-K95).
<sup>k</sup> Assessed using ICD-10 diagnosis codes for diseases of the skin and subcutaneous tissue (L00-L99).
<sup>l</sup> Assessed using ICD-10 diagnosis codes for diseases of the musculoskeletal system and connective tissue (M00-M99).
<sup>m</sup> Assessed using ICD-10 diagnosis codes for diseases of the genitourinary system (N00-N99).
<sup>n</sup> Assessed using ICD-10 diagnosis codes for endocrine, nutritional and metabolic diseases (E00-E89).

The distributions of patient characteristics according to FIASMA medication use are shown in **Table 2**. In the full sample, FIASMA medication use at baseline substantially differed according to all patient characteristics, except for medications according to compassionate use or as part of a clinical trial, and the direction of the associations indicated an older age and greater medical severity of patients receiving FIASMA medication at baseline.

**Table 2.**
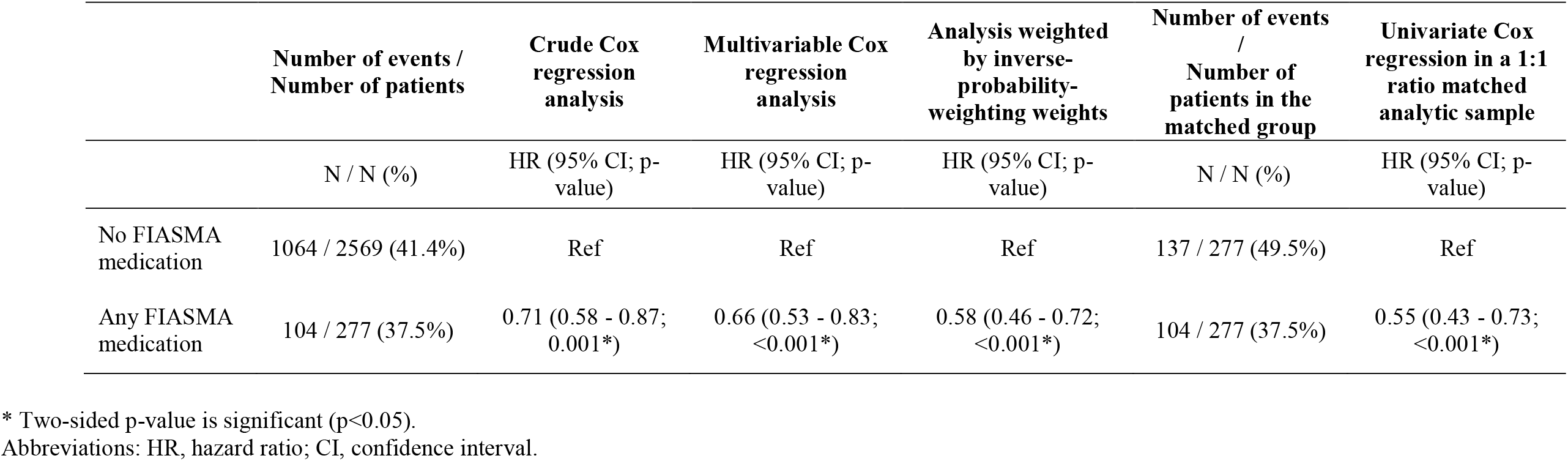
Association between FIASMA medication use at baseline and risk of intubation or death among patients hospitalized for severe COVID-19 (N=2,846).

|  | Number of events /<br>Number of patients | Crude Cox<br>regression<br>analysis | Multivariable Cox<br>regression<br>analysis | Analysis weighted<br>by inverse-probability-weighting weights | Number of events<br>/<br>Number of<br>patients in the<br>matched group | Univariate Cox<br>regression in a 1:1<br>ratio matched<br>analytic sample |
| --- | --- | --- | --- | --- | --- | --- |
|  | N / N (%) | HR (95% CI; p-value) | HR (95% CI; p-value) | HR (95% CI; p-value) | N / N (%) | HR (95% CI; p-value) |
| No FIASMA<br>medication | 1064 / 2569 (41.4%) | Ref | Ref | Ref | 137 / 277 (49.5%) | Ref |
| Any FIASMA<br>medication | 104 / 277 (37.5%) | 0.71 (0.58 - 0.87;<br>0.001*) | 0.66 (0.53 - 0.83;<br><0.001*) | 0.58 (0.46 - 0.72;<br><0.001*) | 104 / 277 (37.5%) | 0.55 (0.43 - 0.73;<br><0.001*) |

\* Two-sided p-value is significant (p<0.05).
Abbreviations: HR, hazard ratio; CI, confidence interval.

In the matched analytic sample and in the full sample after applying the propensity score weights, there were no substantial differences in any characteristic (**Table 2**).

### Study endpoint

The endpoint of intubation or death occurred in 104 patients (37.5%) who received a FIASMA medication at baseline and in 1,060 patients (41.4%) who did not. Both the crude unadjusted analysis (HR=0.71; 95%CI=0.58-0.87; p<0.01) and the primary analysis with inverse probability weighting (HR=0.58; 95% CI=0.46-0.72; p<0.01) showed a significant association between FIASMA medication use at baseline and reduced risk of intubation or death (**Figure 1**; **Table 2**). A post-hoc analysis indicated that we had 80% power in the crude analysis to detect a hazard ratio of at least 0.82 / 1.21.

In sensitivity analyses, the multivariable Cox regression model also showed a significant association (HR=0.66; 95% CI=0.53-0.83; p<0.01), as did the univariate Cox regression model in the matched analytic sample using a 1:1 ratio (HR=0.55; 95% CI=0.43-0.73; p<0.01) (**Table 3**).

Additional exploratory analyses showed that the use of FIASMA nervous system medications, and specifically FIASMA psychoanaleptic medications, was significantly associated with decreased risk of intubation or death across all analyses (**eTable 2**). Using FIASMA cardiovascular system medication, and specifically FIASMA calcium channel blocker medications, was also significantly associated with reduced risk of intubation or death in the primary IPW analysis, multivariable analysis, and the IPW analysis adjusted for unbalanced covariates. Hazard ratios were lower than 1 for most individual FIASMA molecules, but none of them reached statistical significance across all main and sensitivity analyses, except for hydroxyzine and escitalopram (**eTable 2**). Patients receiving any FIASMA medication at baseline, and specifically a FIASMA calcium channel blocker medication, a FIASMA nervous system medication, and specifically a FIASMA psychoanaleptic medication had a significantly reduced risk of intubation or death compared with patients who received paracetamol at baseline (**eTable 3**). Reproducing the main analyses while considering venlafaxine, mirtazapine and citalopram as FIASMA antidepressants did not alter the significance of our results (**eTable 4**). Finally, including in the main analyses all patients with and without clinical severity criteria at baseline did not alter the significance of our results (**eTable 5**).

## Discussion

In this multicenter retrospective observational study involving 2,846 adult patients hospitalized for severe COVID-19, we found that FIASMA medication use at hospital admission was significantly and substantially associated with reduced risk of intubation or death, independently of sociodemographic characteristics and medical comorbidity. This association remained significant in multiple sensitivity analyses. Exploratory analyses suggested that this association was not specific to any particular FIASMA class or medication.

We found that FIASMA medication use among patients hospitalized for severe COVID-19 was significantly and substantially associated with reduced risk of intubation or death, with a 42% risk reduction in the main analysis. This association was not specific to one FIASMA psychotropic class or medication. These findings are in line with prior preclinical^5,10^ and clinical^12–15^ evidence that FIASMA antidepressant medications may substantially prevent cells from being infected with SARS-CoV-2 *in vitro*,^5,10^ and that several FIASMA medications, such as fluoxetine, hydroxyzine and amlodipine at their usual respective antidepressant, antihistaminic, and antihypertensive doses, may reduce mortality among patients hospitalized for COVID-19.^12–15^

Several other mechanisms could be proposed to explain this association besides the involvement of the ASM/ceramide system. First, antiviral effects, i.e. inhibition of viral replication, of FIASMA medications might underlie this relationship, as suggested by *in vitro* studies for fluoxetine,^5^ chlorpromazine,^34^ and amlodipine.^15^ However, inhibition of viral replication was not observed with several other FIASMA medications, including paroxetine and escitalopram.^35^

Second, several FIASMA medications, such as escitalopram or hydroxyzine, have high affinity for Sigma-1 receptors (S1R),^36,37^ which have been suggested to have potential value in regulating inflammation by inhibiting cytokine production in COVID-19.^38^ The S1R has been shown to restrict the endonuclease activity of an Endoplasmic Reticulum (ER) stress sensor called Inositol-Requiring Enzyme1 (IRE1) and to reduce cytokine expression, without inhibiting classical inflammatory pathways.^11,38^ Because several FIASMA medications are S1R agonists in our sample, this mechanism might have overlapped their inhibition effect on ASM. However, when examining the association between the endpoint and several FIASMA medications with low or no affinity for S1R (e.g., amlodipine, paroxetine, duloxetine, and aripiprazole)^36,39–42^, the main results remained statistically significant (**eTable 6**), suggesting that inhibition of ASM could underlie this association independently of S1R.

Finally, this association may be partly mediated by the anti-inflammatory effects of FIASMA medications, which could be explained by inhibition of ASM in endothelial cells and the immune system, and might be independent of Sigma-1 receptors. First, a recent meta-analysis^43^ of studies conducted in individuals with major depressive disorder following antidepressant treatment, mostly including SSRIs, supports that, overall, antidepressants may be associated with decreased plasma levels of 4 of 16 tested inflammatory mediators, including IL-10, TNF-α, CCL-2, and which are associated with COVID-19 severity,^44^ as well as IL-6, which is highly correlated with disease mortality.^44,45^ Second, prior *in vitro* and *in vivo* studies^46–48^ suggest that some antipsychotics may have anti-inflammatory effects via glia activation, but that this activity may not be shared by all antipsychotics. However, this anti-inflammatory effect was observed for both FIASMA antipsychotics (e.g. chlorpromazine) and non-FIASMA ones (e.g., haloperidol and risperidone). If the association between FIASMA psychotropic medication use and reduced risk of intubation or death is confirmed, future studies aiming at disentangling these potentially interrelated mechanisms would be needed.

Our study has several limitations. First, there are two possible major inherent biases in observational studies: unmeasured confounding and confounding by indication. However, in the case of FIASMA medications, including several antidepressants and cardiovascular system medications, confounding by indication may typically result in increased adverse medical outcomes associated with these medications,^49^ not better outcomes as suggested by our findings. We tried to minimize the effects of confounding in several different ways. First, we used an analysis with inverse probability weighting to minimize the effects of confounding by indication,^27,28^ resulting in non-substantial between-group differences in clinical characteristics (all SMD<0.1) in both the IPW primary analysis and the Cox regression analysis in the matched analytic sample. Second, we performed multiple sensitivity analyses, which showed similar results. Finally, although some amount of unmeasured confounding may remain, our analyses adjusted for numerous potential confounders. Other limitations include missing data for some baseline characteristic variables (i.e., 11.5%), which might be explained by the overwhelming of all hospital units during the COVID-19 peak incidence, and different results might have been observed during a lower COVID-19 incidence period. However, imputation of missing data did not alter the significance of our results (data available on request). Second, inflation of type I error might have occurred in exploratory analyses due to multiple testing. Third, data on several FIASMA medications were not available because no patients hospitalized for severe COVID-19 in AP-HP hospitals received them at study baseline during the first epidemic wave. Fourth, this study cannot establish a causal relationship between FIASMA medication use and reduced risk of intubation or death.^50^ Finally, despite the multicenter design, our results may not be generalizable to outpatients or other regions.

In conclusion, in this multicenter observational retrospective study, FIASMA medication use was significantly and substantially associated with reduced risk of intubation or death among adult patients hospitalized for severe COVID-19. These findings suggest the usefulness of the ASM/ceramide system framework in COVID-19 treatment. Double-blind controlled randomized clinical trials of these medications in patients with COVID-19 are needed, starting with FIASMA molecules with the highest *in vitro* inhibition effect on ASM, the highest effect size as observed in our study, and ease of use, including high safety margin, good tolerability, widespread availability, and low cost such that primary care physicians and other providers could prescribe them as soon as onset of symptoms, if their usefulness against COVID-19 was confirmed in RCTs.

## Data Availability

Data can be available at https://eds.aphp.fr// upon request from AP-HP Health Data Warehouse.

## AKNOWLEDGMENT SECTION

### Contributions

NH designed the study, performed statistical analyses, and wrote the first draft of the manuscript. MSR contributed to study design, performed statistical analyses and critically revised the manuscript. FL contributed to study design and critically revised the manuscript for scientific content. RV contributed to statistical analyses and critically revised the manuscript for scientific content. All other authors critically revised the manuscript for scientific content.

### Data Availability Statement

Data can be available at https://eds.aphp.fr// upon request from AP-HP Health Data Warehouse (Entrepôt de Données de Santé (EDS)). The authors did not have special access to the data that other researchers would not have.

### Funding source

This work did not receive any external funding.

### Disclaimer

The information contained in this study is provided for research purpose and should not be used as a substitute or replacement for diagnosis or treatment recommendations or other clinical decisions or judgment. The views presented in this manuscript are those of the authors and should not be construed to represent the views of any of the sponsoring organizations, agencies, or the National Institute on Drug Abuse or any U.S. Government Agency.

## Acknowledgments

The authors thank the EDS APHP Covid consortium integrating the APHP Health Data Warehouse team as well as all the APHP staff and volunteers who contributed to the implementation of the EDS-Covid database and operating solutions for this database. Collaborators of the EDS APHP Covid consortium: Pierre-Yves ANCEL, Alain BAUCHET, Nathanaël BEEKER, Vincent BENOIT, Mélodie BERNAUX, Ali BELLAMINE, Romain BEY, Aurélie BOURMAUD, Stéphane BREANT, Anita BURGUN, Fabrice CARRAT, Charlotte CAUCHETEUX, Julien CHAMP, Sylvie CORMONT, Christel DANIEL, Julien DUBIEL, Catherine DUCLOAS, Loic ESTEVE, Marie FRANK, Nicolas GARCELON, Alexandre GRAMFORT, Nicolas GRIFFON, Olivier GRISEL, Martin GUILBAUD, Claire HASSEN-KHODJA, François HEMERY, Martin HILKA, Anne Sophie JANNOT, Jerome LAMBERT, Richard LAYESE, Judith LEBLANC, Léo LEBOUTER, Guillaume LEMAITRE, Damien LEPROVOST, Ivan LERNER, Kankoe LEVI SALLAH, Aurélien MAIRE, Marie-France MAMZER, Patricia MARTEL, Arthur MENSCH, Thomas MOREAU, Antoine NEURAZ, Nina ORLOVA, Nicolas PARIS, Bastien RANCE, Hélène RAVERA, Antoine ROZES, Elisa SALAMANCA, Arnaud SANDRIN, Patricia SERRE, Xavier TANNIER, Jean-Marc TRELUYER, Damien VAN GYSEL, Gaël VAROQUAUX, Jill Jen VIE, Maxime WACK, Perceval WAJSBURT, Demian WASSERMANN, Eric ZAPLETAL.

## SUPPLEMENTARY INFORMATION

## Supplementary text 1

### FIASMA medications

Several medications, including certain antidepressants (i.e., amitriptyline, clomipramine, desipramine, doxepin, duloxetine, escitalopram, fluoxetine, fluvoxamine, imipramine, lofepramine, maprotiline, nortriptyline, paroxetine, protriptyline, sertraline, and trimipramine), antipsychotics (i.e., aripiprazole, chlorpromazine, chlorprothixene, fluphenazine, flupenthixol, penfluridol, perphenazine, pimozide, promazine, sertindole, thioridazin, trifluoperazine, and triflupromazine), and other medications of the nervous system (cinnarizine, flunarizine), certain antihistaminic medications (astemizole, clemastine, cyproheptadine, desloratadine, hydroxyzine, loratadine, mebhydrolin, pimethixene, promethazine, terfenadine), certain anticholinergic antiparkinson medications (benztropine, biperidene, profenamine), antiprotozoal medications (emetine, quinacrine), calcium channel blockers (amlodipine, bepridil, fendiline, mibefradil, perhexiline), beta blocking agents (carvedilol), antiarrhythmics (amiodarone, aprindine), medications for functional gastrointestinal disorders (alverine, camylofin, dicycloverine, mebeverine), antivertigo medications (cinnarizine, flunarizine), and natural products (conessine, solasodine, tomatidine), a vasodilator (dilazep, suloctidil), a cough suppressant (cloperastine), an antidiarrheal medication (loperamide), an antimycobacterial (clofazimine), an endocrine therapy medication (tamoxifen), and a muscle relaxant (cyclobenzaprine) have shown to *in vitro* inhibit acid sphingomyelinase with varying degrees ^5–9^.

### Supplementary text 2

#### Assessment of variables

We obtained the following data for each patient at the time of the hospitalization: sex; age, which was categorized into 4 classes based on the OpenSAFELY study results ^51^ (i.e. 18-50, 51-70, 71-80, 81+); hospital, which was categorized into 4 classes following the administrative clustering of AP-HP hospitals in Paris and its suburbs based on their geographical location (i.e., AP-HP Centre – Paris University, Henri Mondor University Hospitals and at home hospitalization; AP-HP Nord and Hôpitaux Universitaires Paris Seine-Saint-Denis; AP-HP Paris Saclay University; and AP-HP Sorbonne University); obesity, which was defined as having a body mass index higher than 30 kg/m^2^ or an International Statistical Classification of Diseases and Related Health Problems (ICD-10) diagnosis code for obesity (E66.0, E66.1, E66.2, E66.8, E66.9); self-reported current smoking status; and any medication prescribed according to compassionate use or as part of a clinical trial (e.g. hydroxychloroquine, azithromycin, remdesivir, tocilizumab, sarilumab, or dexamethasone). To take into account possible confounding by indication bias for FIASMA medications, we recorded whether patients had any current diagnosis, based on ICD-10 diagnosis codes recorded during the visit, of neoplasms and diseases of the blood (C00-D89); mental disorders (F01-F99); diseases of the nervous system (G00-G99); cardiovascular disorders (I00-I99); respiratory disorders (J00-J99); digestive disorders (K00-K95); dermatological disorders (L00-L99); diseases of the musculoskeletal system (M00-M99); diseases of the genitourinary system (N00-N99); endocrine disorders (E00-E89); and eye-ear-nose-throat disorders (H00-H95).

All medical notes and prescriptions are computerized in Greater Paris University hospitals. Medications including their dose, frequency, date, and mode of administration were identified from medication administration data or scanned hand-written medical prescriptions, through two deep learning models based on BERT contextual embeddings ^23^, one for the medications and another for their mode of administration. The model was trained on the APmed corpus ^24^, a previously annotated dataset for this task. Extracted medications names were then normalized to the Anatomical Therapeutic Chemical (ATC) terminology using approximate string matching.

**eTable 1.** Associations of baseline clinical characteristics with the composite endpoint of intubation or death in the cohort of adult patients who had been admitted to AP-HP hospitals for severe COVID-19 (N=2,846).

|  | Full sample<br>(N=2,846) | With the<br>endpoint<br>(N=1,168) | Without the<br>endpoint<br>(N=1,678) | Crude analysis | Multivariable analysis |  |
| --- | --- | --- | --- | --- | --- | --- |
|  | N (%) | N (%) | N (%) | HR (95% CI;<br>p-value) | HR (95% CI; p-value) | Collinearity<br>diagnostic<br>(variance<br>inflation factor) |
| Age |  |  |  |  |  | 1,04 |
| 18 to 50 years | 522 (18.3) | 137 (26.2) | 385 (73.8) | Ref. | Ref. |  |
| 51 to 70 years | 1115 (39.2) | 438 (39.3) | 677 (60.7) | 1.27 (1.05 -<br>1.54; 0.013*) | 1,07 (0,86 -<br>1,33; 0,567) |  |
| 71 to 80 years | 520 (18.3) | 242 (46.5) | 278 (53.5) | 1.41 (1.14 -<br>1.74; 0.001*) | 1,08 (0,84 -<br>1,38; 0,565) |  |
| More than 80 years | 689 (24.2) | 351 (50.9) | 338 (49.1) | 1.37 (1.13 -<br>1.67; 0.002*) | 1,16 (0,90 -<br>1,50; 0,250) |  |
| Sex |  |  |  |  |  | 1,05 |
| Women | 1064 (37.4) | 375 (35.2) | 689 (64.8) | Ref. | Ref. |  |
| Men | 1782 (62.6) | 793 (44.5) | 989 (55.5) | 1.28 (1.13 -<br>1.45; <0.001*) | 1,50 (1,26 -<br>1,79; <0,001*) |  |
| Hospital |  |  |  |  |  | 1,02 |
| AP-HP Centre - Paris University,<br>Henri Mondor University Hospitals and<br>at home hospitalization | 722 (25.4) | 349 (48.3) | 373 (51.7) | Ref. | Ref. |  |
| *AP-HP Nord and Hôpitaux*<br>*Universitaires Paris Seine-Saint-Denis* | 889 (31.2) | 313 (35.2) | 576 (64.8) | 0.87 (0.74 -<br>1.01; 0.067) | 1,05 (0,87 -<br>1,27; 0,590) |  |
| *AP-HP Paris Saclay University* | 624 (21.9) | 245 (39.3) | 379 (60.7) | 0.77 (0.65 -<br>0.91; 0.002*) | 0,77 (0,63 -<br>0,93; 0,009*) |  |
| *AP-HP Sorbonne University* | 611 (21.5) | 261 (42.7) | 350 (57.3) | 0.94 (0.80 -<br>1.10; 0.452) | 1,00 (0,83 -<br>1,22; 0,963) |  |
| Obesity <sup>a</sup> |  |  |  |  |  | 1,08 |
| *Yes* | 582 (20.4) | 293 (50.3) | 289 (49.7) | 1.34 (1.17 -<br>1.53; <0.001*) | 1,31 (1,08 -<br>1,59; 0,005*) |  |
| *No* | 2264 (79.6) | 875 (38.6) | 1389 (61.4) | Ref. | Ref. |  |
| Smoking <sup>b</sup> |  |  |  |  |  | 1,02 |
| *Yes* | 356 (12.5) | 161 (45.2) | 195 (54.8) | 0.94 (0.80 -<br>1.11; 0.463) | 0,90 (0,71 -<br>1,14; 0,389) |  |
| *No* | 2490 (87.5) | 1007 (40.4) | 1483 (59.6) | Ref. | Ref. |  |
| Medication according to compassionate<br>use or as part of a clinical trial <sup>c</sup> |  |  |  |  |  | 1,06 |
| *Yes* | 806 (28.3) | 343 (42.6) | 463 (57.4) | 0.95 (0.84 -<br>1.08; 0.450) | 0,80 (0,69 -<br>0,93; 0,003*) |  |
| *No* | 2040 (71.7) | 825 (40.4) | 1215 (59.6) | Ref. | Ref. |  |
| Other infectious diseases <sup>d</sup> |  |  |  |  |  | 1,08 |
| *Yes* | 356 (12.5) | 221 (62.1) | 135 (37.9) | 1.71 (1.48 -<br>1.98; <0.001*) | 1,30 (1,08 -<br>1,56; 0,006*) |  |
| *No* | 2490 (87.5) | 947 (38.0) | 1543 (62.0) | Ref. | Ref. |  |
| Neoplasms and diseases of the blood <sup>e</sup> |  |  |  |  |  | 1,07 |
| *Yes* | 339 (11.9) | 177 (52.2) | 162 (47.8) | 1.22 (1.04 -<br>1.43; 0.017*) | 0,79 (0,64 -<br>0,98; 0,035*) |  |
| *No* | 2507 (88.1) | 991 (39.5) | 1516 (60.5) | Ref. | Ref. |  |
| Mental disorders <sup>f</sup> |  |  |  |  |  | 1,09 |
| *Yes* | 340 (11.9) | 198 (58.2) | 142 (41.8) | 1.37 (1.17 -<br>1.59; <0.001*) | 1,10 (0,92 -<br>1,32; 0,291) |  |
| *No* | 2506 (88.1) | 970 (38.7) | 1536 (61.3) | Ref. | Ref. |  |
| Diseases of the nervous system <sup>g</sup> |  |  |  |  |  | 1,08 |
| *Yes* | 292 (10.3) | 174 (59.6) | 118 (40.4) | 1.54 (1.31 - 1.81; <0.001*) | 1,05 (0,86 - 1,27; 0,639) |  |
| *No* | 2554 (89.7) | 994 (38.9) | 1560 (61.1) | Ref. | Ref. |  |
| Cardiovascular disorders <sup>h</sup> |  |  |  |  |  | 1,24 |
| *Yes* | 1020 (35.8) | 557 (54.6) | 463 (45.4) | 1.78 (1.59 - 2.00; <0.001*) | 1,29 (1,09 - 1,53; 0,003*) |  |
| *No* | 1826 (64.2) | 611 (33.5) | 1215 (66.5) | Ref. | Ref. |  |
| Respiratory disorders <sup>i</sup> |  |  |  |  |  | 1,25 |
| *Yes* | 1704 (59.9) | 828 (48.6) | 876 (51.4) | 1.90 (1.67 - 2.16; <0.001*) | 1,38 (1,10 - 1,72; 0,005*) |  |
| *No* | 1142 (40.1) | 340 (29.8) | 802 (70.2) | Ref. | Ref. |  |
| Digestive disorders <sup>j</sup> |  |  |  |  |  | 1,05 |
| *Yes* | 234 (8.2) | 121 (51.7) | 113 (48.3) | 1.22 (1.01 - 1.47; 0.038*) | 1,03 (0,84 - 1,26; 0,769) |  |
| *No* | 2612 (91.8) | 1047 (40.1) | 1565 (59.9) | Ref. | Ref. |  |
| Dermatological disorders <sup>k</sup> |  |  |  |  |  | 1,02 |
| *Yes* | 71 (2.5) | 44 (62.0) | 27 (38.0) | 1.38 (1.02 - 1.86; 0.036*) | 1,07 (0,75 - 1,52; 0,721) |  |
| *No* | 2775 (97.5) | 1124 (40.5) | 1651 (59.5) | Ref. | Ref. |  |
| Diseases of the musculoskeletal system <sup>l</sup> |  |  |  |  |  | 1,04 |
| *Yes* | 143 (5.0) | 67 (46.9) | 76 (53.1) | 1.02 (0.80 - 1.30; 0.890) | 0,94 (0,73 - 1,22; 0,655) |  |
| *No* | 2703 (95.0) | 1101 (40.7) | 1602 (59.3) | Ref. | Ref. |  |
| Diseases of the genitourinary system <sup>m</sup> |  |  |  |  |  | 1,10 |
| *Yes* | 429 (15.1) | 288 (67.1) | 141 (32.9) | 1.93 (1.69 - 2.20; <0.001*) | 1,25 (1,01 - 1,56; 0,043*) |  |
| *No* | 2417 (84.9) | 880 (36.4) | 1537 (63.6) | Ref. | Ref. |  |
| Endocrine disorders <sup>n</sup> |  |  |  |  |  | 1,27 |
| *Yes* | 1042 (36.6) | 542 (52.0) | 500 (48.0) | 1.56 (1.39 - 1.75; <0.001*) | 1,12 (0,95 - 1,33; 0,173) |  |
| *No* | 1804 (63.4) | 626 (34.7) | 1178 (65.3) | Ref. | Ref. |  |
| Eye-Ear-Nose-Throat disorders <sup>o</sup> |  |  |  |  |  | 1,03 |
| *Yes* | 59 (2.1) | 31 (52.5) | 28 (47.5) | 1.18 (0.82 - 1.68; 0.370) | 0,97 (0,69 - 1,35; 0,840) |  |
| *No* | 2787 (97.9) | 1137 (40.8) | 1650 (59.2) | Ref. | Ref. |  |

<sup>a</sup> Defined as having a body-mass index higher than 30 kg/m<sup>2</sup> or an International Statistical Classification of Diseases and Related Health Problems (ICD-10) diagnosis code for obesity (E66.0, E66.1, E66.2, E66.8, E66.9).
<sup>b</sup> Current Smoking status was self-reported.
<sup>c</sup> Any medication prescribed as part of a clinical trial or according to compassionate use (e.g., hydroxychloroquine, azithromycin, remdesivir, tocilizumab, sarilumab or dexamethasone).
<sup>d</sup> Assessed using ICD-10 diagnosis codes for certain infectious and parasitic diseases (A00-B99).
<sup>e</sup> Assessed using ICD-10 diagnosis codes for neoplasms (C00-D49) and diseases of the blood and blood-forming organs and certain disorders involving the immune mechanism (D50-D89).
<sup>f</sup> Assessed using ICD-10 diagnosis codes for mental, behavioural and neurodevelopmental disorders (F01-F99).
<sup>g</sup> Assessed using ICD-10 diagnosis codes for diseases of the nervous system (G00-G99).
<sup>h</sup> Assessed using ICD-10 diagnosis codes for diseases of the circulatory system (I00-I99).
<sup>i</sup> Assessed using ICD-10 diagnosis codes for diseases of the respiratory system (J00-J99).
<sup>j</sup> Assessed using ICD-10 diagnosis codes for diseases of the digestive system (K00-K95).
<sup>k</sup> Assessed using ICD-10 diagnosis codes for diseases of the skin and subcutaneous tissue (L00-L99).
<sup>l</sup> Assessed using ICD-10 diagnosis codes for diseases of the musculoskeletal system and connective tissue (M00-M99).
<sup>m</sup> Assessed using ICD-10 diagnosis codes for diseases of the genitourinary system (N00-N99).
<sup>n</sup> Assessed using ICD-10 diagnosis codes for endocrine, nutritional and metabolic diseases (E00-E89).

**eTable 2.**
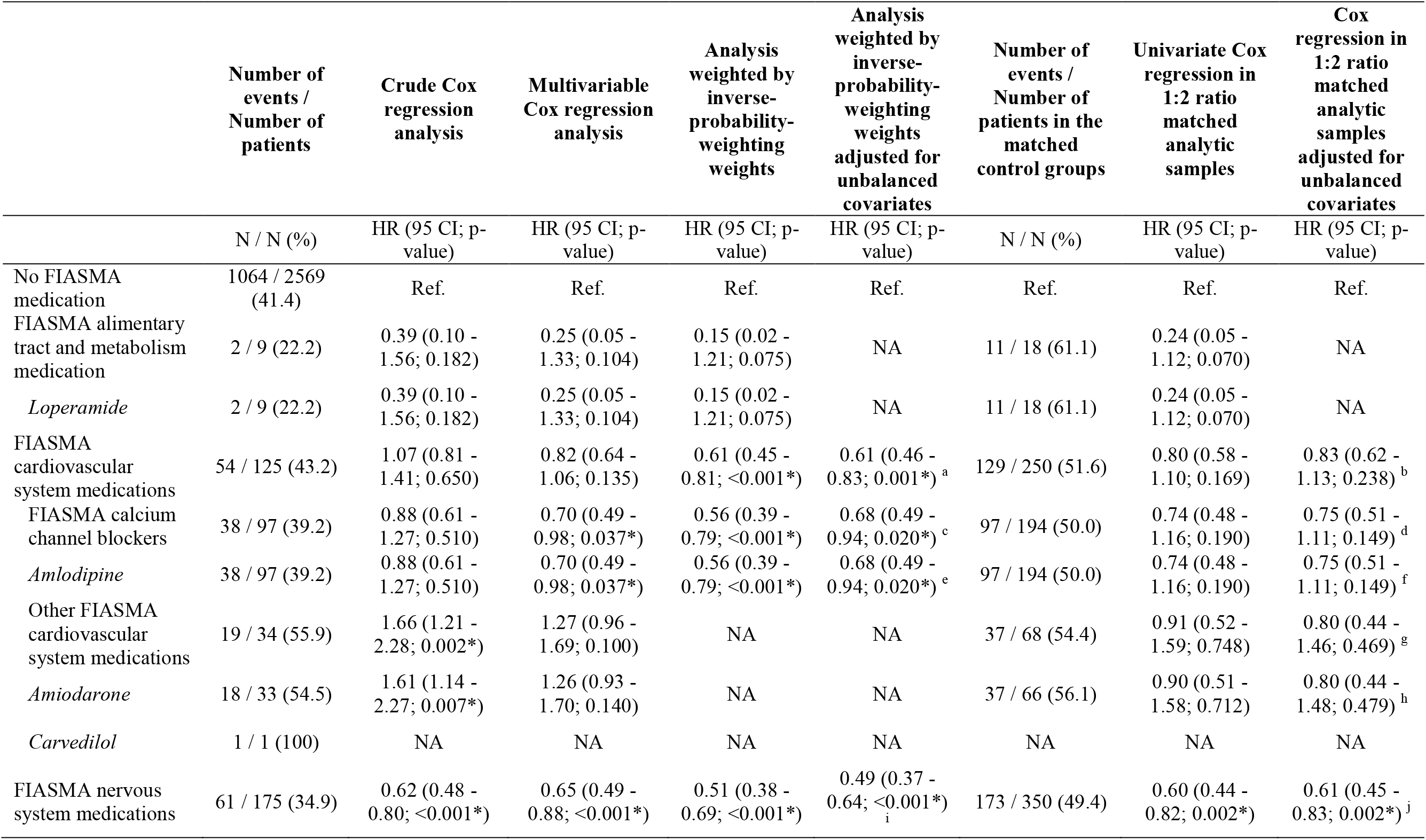
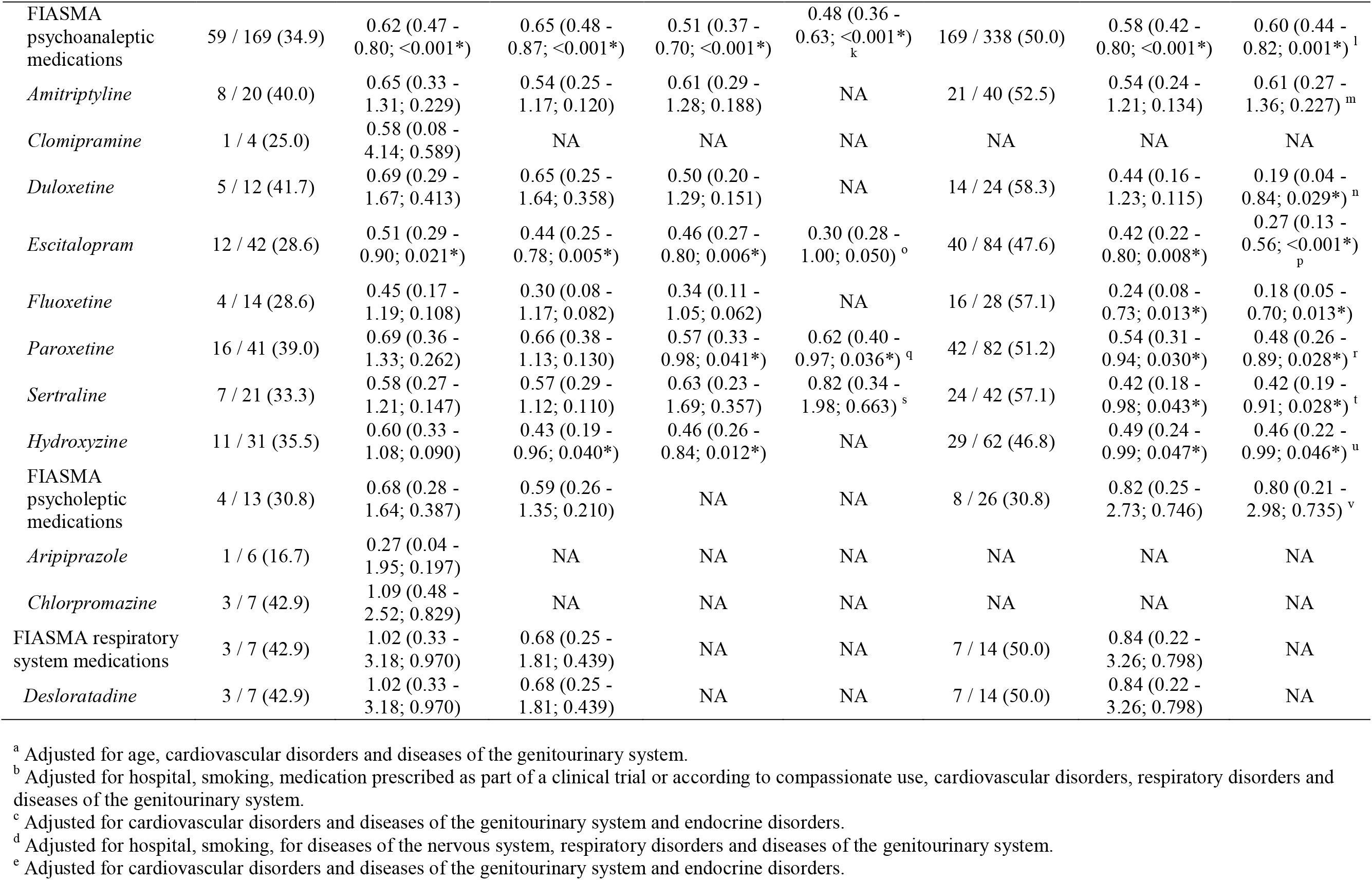
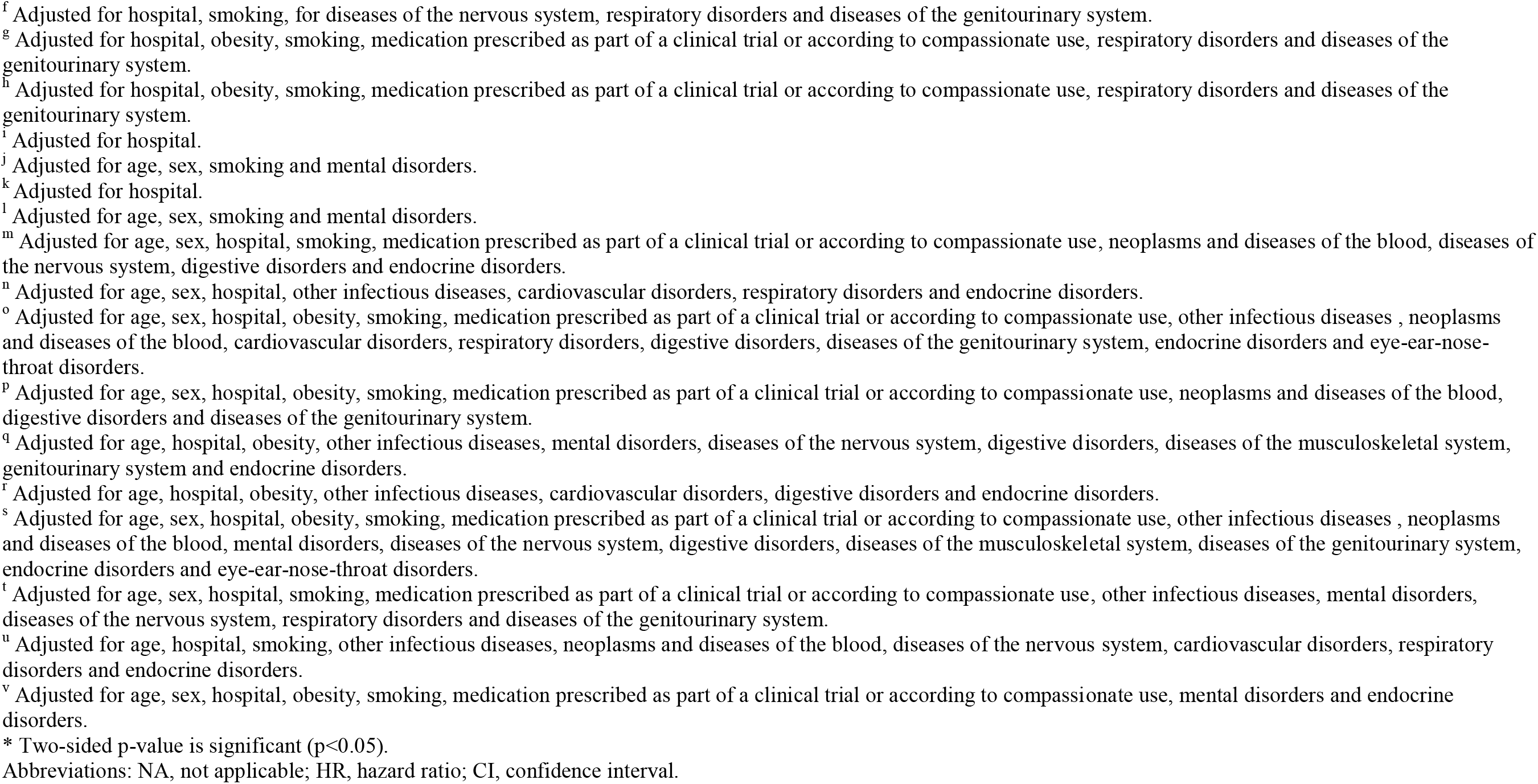
Association of each FIASMA class and molecule prescribed at baseline with the composite endpoint of intubation or death among patients hospitalized for severe COVID-19.

**eTable 3.**
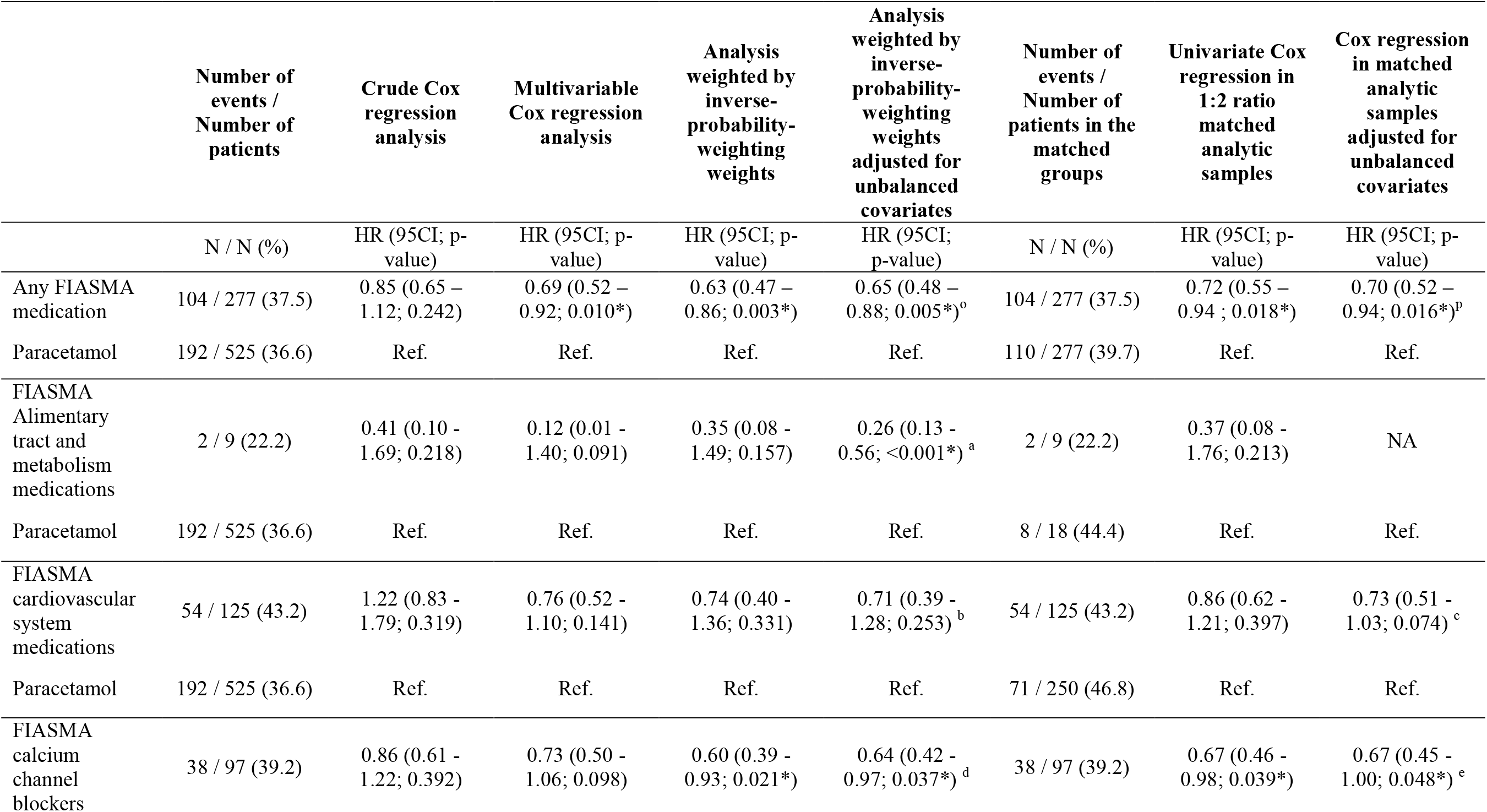
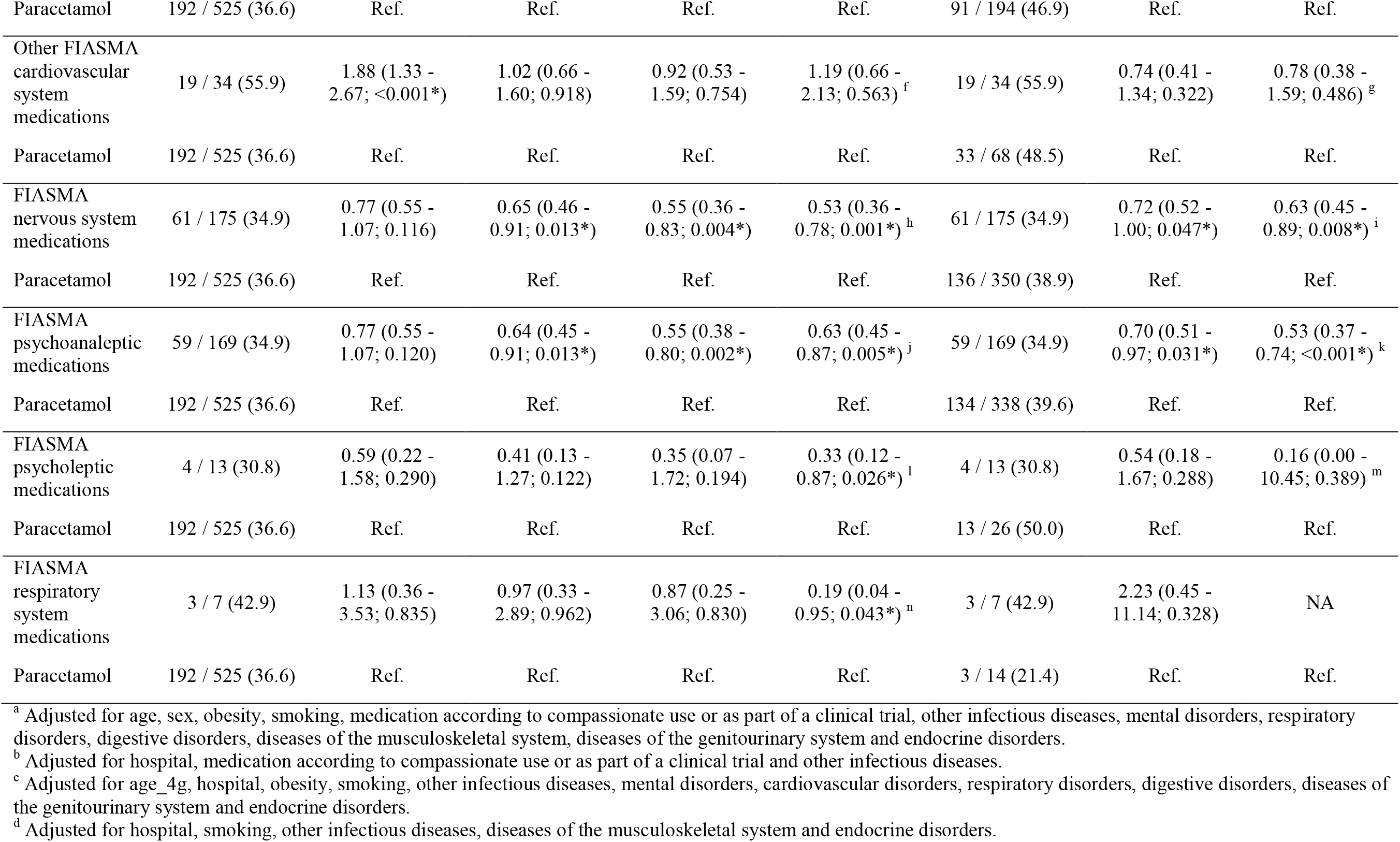
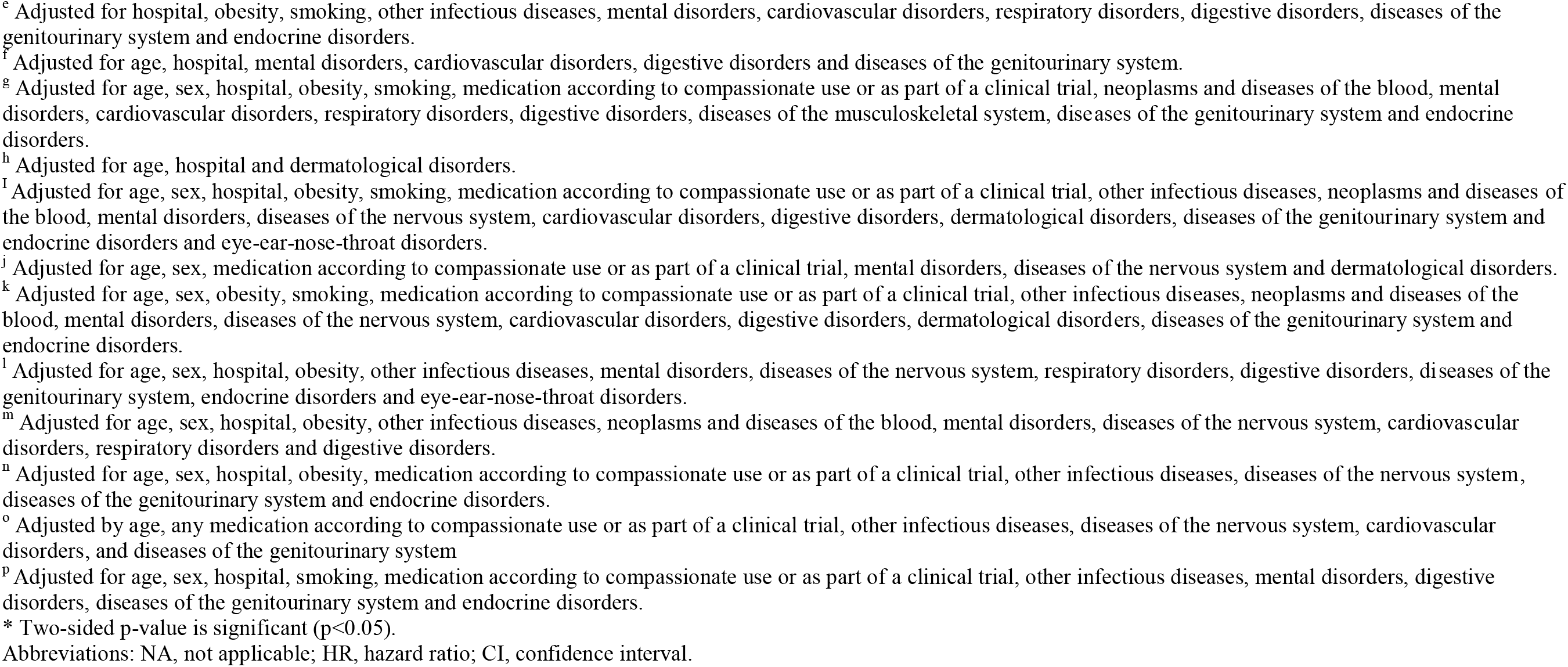
Association between FIASMA medication use at baseline and the endpoint of intubation or death as compared with paracetamol use at baseline among patients hospitalized for severe COVID-19.

**eTable 4.** Association between FIASMA medication use at baseline and the composite endpoint of intubation or death among patients hospitalized for severe COVID-19, while considering venlafaxine, citalopram, and mirtazapine as FIASMAs (N=2,846).

|  | Number of events<br>/ Number of<br>patients | Crude Cox<br>regression<br>analysis | Multivariable<br>Cox regression<br>analysis | Analysis weighted<br>by inverse-probability-weighting weights | Analysis weighted<br>by inverse-probability-weighting weights<br>adjusted for<br>unbalanced<br>covariates <sup>a</sup> | Number of<br>events /<br>Number of<br>patients in the<br>matched<br>group | Univariate Cox<br>regression in a<br>1:1 ratio<br>matched<br>analytic sample | Cox<br>regression in<br>a 1:1 ratio<br>matched<br>analytic<br>sample<br>adjusted for<br>unbalanced<br>covariates <sup>b</sup> |
| --- | --- | --- | --- | --- | --- | --- | --- | --- |
|  | N / N (%) | HR (95 CI; p-value) | HR (95 CI; p-value) | HR (95 CI; p-value) | HR (95 CI; p-value) | N / N (%) | HR (95 CI; p-value) | HR (95CI; p-value) |
| No<br>FIASMA<br>medication | 1,059 / 2,559<br>(41.4) | Ref | Ref | Ref | Ref | 141 / 287<br>(49.1) | Ref | Ref |
| Any<br>FIASMA<br>medication | 109 / 287 (38.0) | 0.73 (0.60 –<br>0.89; 0.002*) | 0.67 (0.54 – 0.84;<br><0.001*) | 0.55 (0.45 – 0.68;<br><0.001*) | 0.55 (0.45 – 0.68;<br><0.001*) | 109 / 287<br>(38.0) | 0.59 (0.45 –<br>0.78; <0.001*) | 0.60 (0.45 –<br>0.79; <0.001*) |

<sup>a</sup> All covariates were balanced.
<sup>b</sup> Adjusted for hospital.
\* Two-sided p-value is significant (p<0.05).
Abbreviations: HR, hazard ratio; CI, confidence interval.

**eTable 5.**
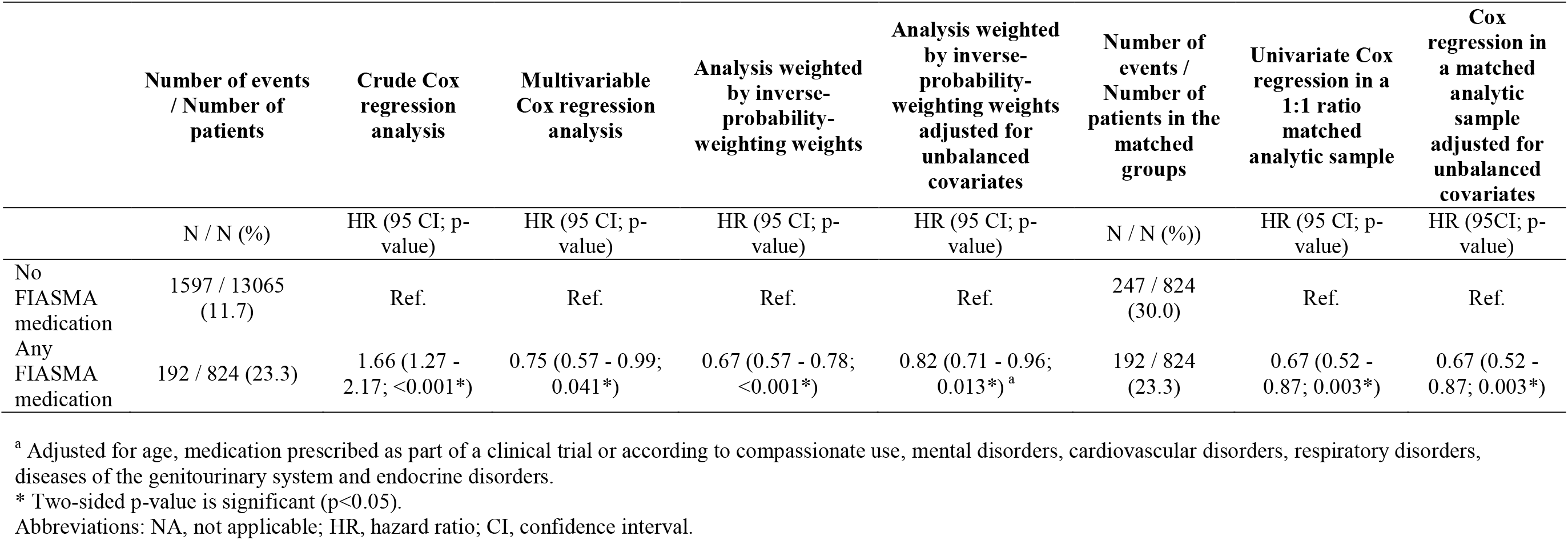
Association between FIASMA medication use at baseline and the composite endpoint of intubation or death among patients hospitalized for COVID-19 with or without clinical severity of COVID-19 at baseline (N=14,429).

**eTable 6.** Association between FIASMA medications with low or no affinity for Sigma-1-receptors (i.e., amlodipine, paroxetine, duloxetine, aripiprazole) and the composite endpoint of intubation or death among adult patients hospitalized for severe COVID-19.

|  | Number of events /<br>Number of<br>patients | Crude Cox<br>regression analysis | Multivariable<br>Cox regression<br>analysis | Analysis weighted<br>by inverse-probability-weighting weights | Analysis weighted<br>by inverse-probability-weighting weights<br>adjusted for<br>unbalanced<br>covariates <sup>a</sup> | Number of<br>events /<br>Number of<br>patients in the<br>matched<br>groups | Univariate Cox<br>regression in a<br>1:1 ratio<br>matched analytic<br>sample | Cox regression<br>in a 1:1 ratio<br>matched analytic<br>sample adjusted<br>for unbalanced<br>covariates <sup>b</sup> |
| --- | --- | --- | --- | --- | --- | --- | --- | --- |
|  | N / N (%) | HR (95% CI; p-value) | HR (95% CI; p-value) | HR (95% CI; p-value) | HR (95% CI; p-value) | N / N (%) | HR (95% CI; p-value) | HR (95% CI; p-value) |
| No<br>FIASMA<br>medication | 1064 / 2569 (41.4) | Ref. | Ref. | Ref. | Ref. | 77 / 145 (53.1) | Ref. | Ref. |
| FIASMA<br>medications<br>with low or<br>no affinity<br>for Sigma-1-receptors | 55 / 145 (37.9) | 0.76 (0.555 – 1.06;<br>0.108) | 0.65 (0.49 – 0.87;<br>0.004*) | 0.56 (0.42 – 0.74;<br><0.001*) | 0.65 (0.49 – 0.85;<br>0.002*) | 55 / 145 (37.9) | 0.47 (0.31 – 0.69;<br><0.001*) | 0.46 (0.32 – 0.68;<br><0.001*) |

\* Two-sided p-value is significant (p<0.05).
<sup>a</sup> Adjusted for cardiovascular disorders, diseases of the genitourinary system, endocrine disorders, and eye-ear-nose-throat disorders.
<sup>b</sup> Adjusted for hospital, mental disorders, and respiratory disorders.
Abbreviations: HR, hazard ratio; CI, confidence interval.

